# No Evidence for Increased Platelet Reactivity with Xylitol or Erythritol Sweeteners in Healthy Humans

**DOI:** 10.64898/2026.09.25.26364060

**Authors:** Bettina K. Wölnerhanssen, Anita Altstädt, Emilie Flad, Mariantonietta Tripodo, Tanja Knopp, Jürgen Drewe, Stefan Gaugler, Anne Angelillo-Scherrer, Anne Christin Meyer-Gerspach

**Author notes:** Corresponding author: Bettina K. Wölnerhanssen, Metabolic and Gastroenterological Research St. Clara Research Ltd., 4002 Basel, Switzerland. Shared first author (Bettina K. Wölnerhanssen, Anita Altstädt, Emilie Flad). Shared last author (Anne Angelillo-Scherrer, Anne Christin Meyer-Gerspach).

## Abstract

**Background:** Erythritol and xylitol, naturally present in fruits and vegetables, are widely used as alternative sweeteners with minimal glycemic and insulinemic effects. Both are also produced endogenously, with elevated plasma concentrations reported in metabolic disorders and cardiovascular disease. Recently, two interventional studies reported enhanced platelet aggregation after acute oral intake of erythritol or xylitol, raising concerns about potential prothrombotic effects. We investigated whether acute exposure to erythritol or xylitol increases platelet reactivity in healthy individuals.

**Methods:** We conducted a randomized, single-blind, crossover study in 14 healthy participants who received erythritol (50 g), xylitol (33.5 g), or water. Platelet aggregation was assessed *ex vivo*, together with circulating markers of platelet activation (P-selectin), endothelial activation (sVCAM-1), and coagulation (D-dimer), as well as plasma erythritol and xylitol concentrations. Complementary *in vitro* experiments were performed using platelet-rich plasma from 11 healthy participants exposed to increasing concentrations of erythritol or xylitol, followed by assessment of platelet aggregation.

**Results:** Despite substantial increases in circulating erythritol and xylitol concentrations following oral intake, neither erythritol nor xylitol increased *ex vivo* platelet aggregation compared with water. Consistently, no relevant changes were observed in P-selectin, sVCAM-1, or D-dimer concentrations. Direct *in vitro* exposure of platelet-rich plasma to increasing concentrations of erythritol or xylitol likewise did not enhance platelet aggregation.

**Conclusions:** In healthy individuals, acute oral intake of erythritol or xylitol did not increase platelet aggregation or circulating markers of platelet activation, endothelial activation, or coagulation. These findings were supported by *in vitro* experiments showing no direct effect of erythritol or xylitol on platelet aggregation. Our results do not support an acute platelet-activating effect of erythritol or xylitol under the conditions studied.

**Registration:** URL: https://clinicaltrials.gov/study/NCT04966299; Unique identifier: NCT04966299.

**Graphical Abstract:** 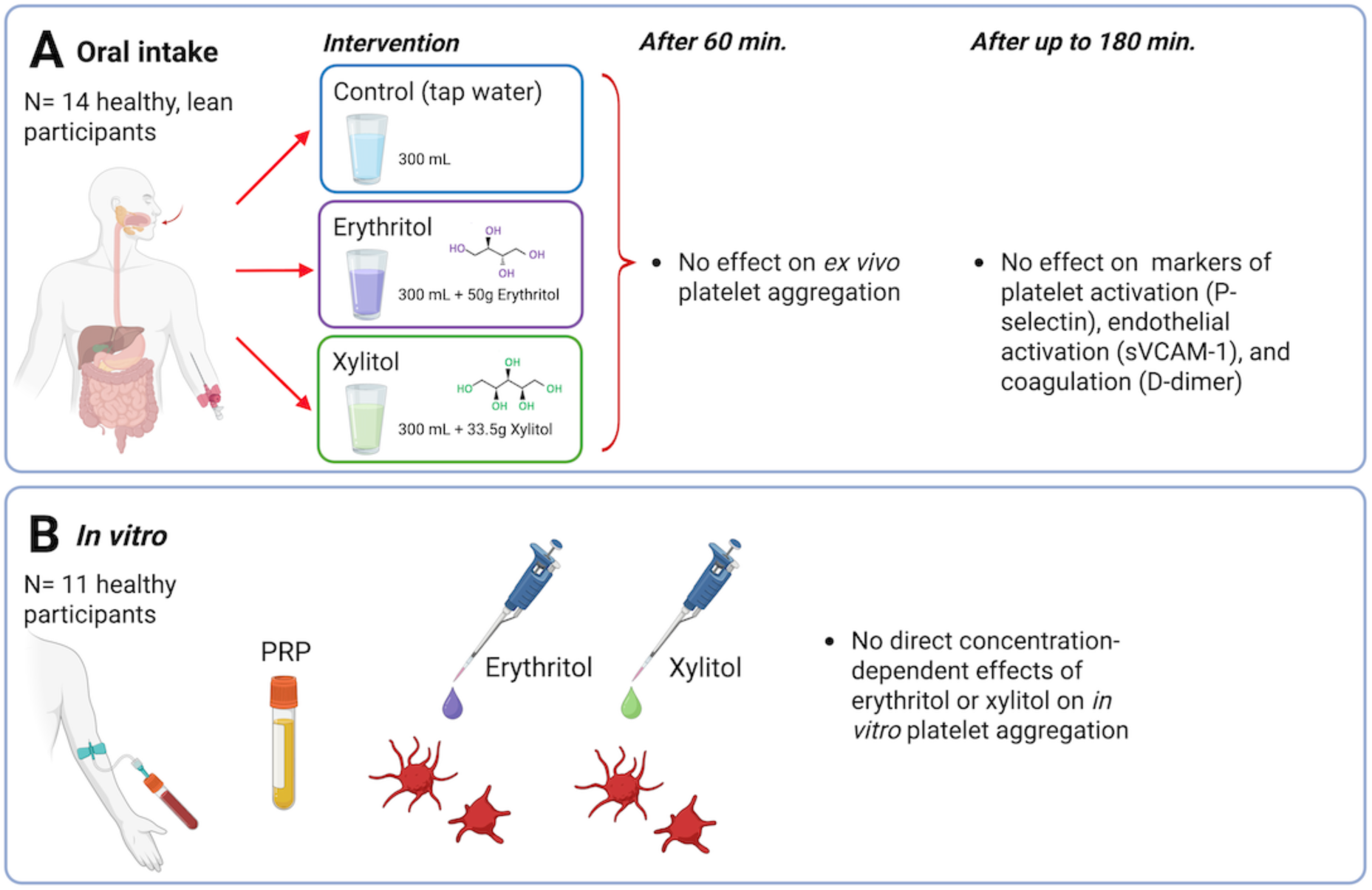

Effect of erythritol and xylitol versus water (control) on platelet reactivity. TRAP-6, thrombin receptor-activating peptide; ADP, adenosine diphosphate; PRP, platelet-rich plasma.

**What Are the Clinical Implications?:** Recent human intervention studies have raised concerns that acute erythritol or xylitol intake may enhance platelet reactivity and promote a prothrombotic state. In the present randomized crossover study of healthy adults, neither erythritol nor xylitol increased platelet aggregation or circulating markers of platelet activation, endothelial activation, or coagulation. These findings do not support an acute prothrombotic effect of erythritol or xylitol under the conditions studied and provide reassurance regarding their immediate effects on platelet function in healthy individuals. However, they do not establish long-term cardiovascular safety. Larger, longer-term studies, particularly in individuals at increased cardiovascular risk, are needed to determine the clinical relevance of chronic erythritol and xylitol exposure.

## Introduction

High intake of dietary sugars is associated with a broad range of metabolic disturbances, including weight gain, insulin resistance, type 2 diabetes mellitus (T2DM), dyslipidemia, hypertension, inflammation, and non-alcoholic fatty liver disease, all of which collectively increase cardiovascular risk ^1^.

Both acute and chronic hyperglycemia together with insulin resistance promote a prothrombotic state in individuals with and without T2DM, characterized by altered platelet number and function, increased platelet sensitivity and activation of the coagulation cascade ^2–4^. Accordingly, limiting dietary sugar intake is recommended for the general population, including people with T2DM, to reduce cardiovascular risk.

Erythritol and xylitol, naturally present in fruits and vegetables, have gained popularity as alternative sweeteners as they provide sweetness with minimal glycemic and insulinemic effects ^5,6^. Both erythritol and xylitol are also produced endogenously in small amounts *via* the pentose phosphate and glucuronic acid pathways ^7–9^. Elevated endogenous polyol production - including erythritol, xylitol, arabitol, and ribitol - has been observed in metabolic disorders including obesity, T2DM, and cardiovascular disease (CVD) ^7,10–12^. Two recent observational studies have reported associations between elevated plasma erythritol or xylitol concentrations and increased cardiovascular risk ^11,13^. However, these studies could not distinguish between endogenous production and exogenous intake of erythritol or xylitol, nor infer causality. The mechanisms underlying elevated circulating polyol concentrations remain incompletely understood.

More recently, two interventional studies in healthy participants suggested that acute oral intake of erythritol or xylitol may enhance platelet aggregation ^13,14^. These findings raised concerns about potential prothrombotic effects of erythritol and xylitol and their implications for cardiovascular health, warranting further investigation. We therefore carried out a two-part study to examine whether erythritol or xylitol influences platelet reactivity. Part A involved acute oral intake of erythritol or xylitol in healthy participants and assessed (i) *ex vivo* platelet aggregation, (ii) markers of platelet activation (P-selectin), endothelial activation (sVCAM-1), and coagulation (D-dimer), as well as (iii) plasma erythritol and xylitol concentrations. Part B consisted of *in vitro* experiments investigating the direct effects of erythritol or xylitol on platelet aggregation. We hypothesized that neither erythritol nor xylitol would enhance platelet aggregation *ex vivo* or *in vitro*.

## Methods

### Ethics and Registration

The study (Part A and B) was approved by the local ethics committee in Basel (Ethikkommission Nordwest- und Zentralschweiz (EKNZ): 2021-00626) and conducted in accordance with the current version of the Declaration of Helsinki (2024), the International Council for Harmonisation Good Clinical Practice (ICH-GCP) guidelines, and all applicable national legal and regulatory requirements. All participants provided written informed consent before enrollment. The study was registered at ClinicalTrials.gov (NCT04966299).

### Part A. Effects of Acute Oral Intake of Erythritol or Xylitol

#### Study Design and Participants

This was a randomized, single-blind, crossover study. Each participant completed three study visits separated by ≥1 week. Twenty healthy, non-smoking adults (age: 18–55 years; body mass index (BMI): 19.0–24.9 kg/m^2^) were enrolled. Key exclusion criteria included acute or chronic illness, pregnancy, substance abuse, smoking, regular intake (>1x per week) of erythritol or xylitol, known impairment of blood coagulation or platelet function, use of medication affecting coagulation or platelet function, or participation in another clinical study with investigational drugs within 30 days prior to or during the study. Participants were instructed to abstain from any medication affecting blood coagulation for at least two weeks before each study visit, avoid vigorous physical activity for 24 hours before each visit, and fast from 8:00 pm the evening before each visit.

At each visit, participants consumed one of three treatments in randomized order within 2 min:

i. Erythritol: 50 g dissolved in 300 mL water
ii. Xylitol: 33.5 g dissolved in 300 mL water
iii. Water control: 300 mL plain water

The allocation sequence was computer-generated. Erythritol and xylitol were purchased from Schweizer Edelzucker AG (St. Gallen, Switzerland). All treatments were freshly prepared at room temperature each morning. Participants and laboratory personnel were blinded to the sweetener condition; complete blinding was not possible for the water condition resulting in partial blinding at the participant level. Blood pressure and heart rate were measured at the beginning and end of each visit.

#### Blood Collection and Processing

For *ex vivo* platelet aggregation, blood was drawn with a 19-gauge butterfly needle at t = −60 min and 60 min. To minimize potential platelet activation related to venipuncture, the first 2.6 mL were discarded, after which blood was collected into citrate tubes (9NC, 0.129 mol/L, 3.8 % buffered) for platelet function assay (PFA) ^15^. Immediately after collection, the tubes were gently inverted several times to ensure adequate mixing of blood with the citrate anticoagulant while avoiding vigorous agitation that could induce platelet activation. The samples were then kept undisturbed at room temperature for 15 min before further processing, in accordance with our standardized pre-analytical procedure for platelet function testing. This resting period allowed standardized handling of the samples after venipuncture and before centrifugation ^16^.

Platelet-rich plasma (PRP) was subsequently prepared by centrifugation at 200 × g for 10 min without brake. Platelet-poor plasma (PPP) was prepared by centrifugation at 1500 × g for 15 min. PRP and PPP were used for aggregometry within 60 min of blood collection. For assessment of markers of platelet activation (P-selectin), endothelial activation (sVCAM-1), and coagulation (D-dimer), and plasma erythritol and xylitol concentrations, a forearm venous cannula was inserted and blood was collected at t = −60, −1, 30, 60, 120, and 180 min; additional samples for erythritol and xylitol measurements were taken at 24 h and 48 h. Ethylenediaminetetraacetic acid (EDTA) tubes (6 µmol/L blood) were used for P-selectin, sVCAM-1, and erythritol and xylitol measurements, and citrate tubes (0.106 mol/L blood) for D-dimer. Samples were immediately placed on ice, centrifuged (EDTA: 4 °C, 3000 × g, 10 min; citrate: 4 °C, 2000 × g, 10 min), and stored as plasma aliquots at −80 °C until analysis.

#### Laboratory Analyses

*Ex vivo* platelet aggregation was measured turbidimetrically using an optical aggregometer (APACT 4004, Coultronics). PRP was adjusted to a platelet concentration of 250 × 10⁹/L using autologous PPP. Aggregation was induced with ADP (2 µM) or TRAP-6 (5 µM and 10 µM). The agonist concentrations were selected based on the dose-response experiments reported by Witkowski *et al.* ^13,14^. Specifically, 2 µM ADP was chosen as a submaximal concentration at which erythritol-induced enhancement of platelet responsiveness was previously demonstrated. For TRAP-6, 5 and 10 µM were selected to assess platelet aggregation under submaximal and stronger stimulation, respectively. Thus, rather than reproducing a complete dose-response curve, we selected concentrations that allowed us to assess erythritol-related changes in platelet responsiveness across different levels of platelet activation. Maximum aggregation, inclination, lag phase, and de-aggregation were quantified to capture complementary aspects of platelet activation kinetics and aggregate stability.

Plasma concentrations of P-selectin and sVCAM-1 were measured using commercially available DuoSet enzyme-linked immunosorbent assay (ELISA) kits (R&D Systems, Minneapolis, MN, USA; Catalog #DY137 and #DY809, respectively). D-dimer concentrations were assessed using an automated, latex-enhanced turbidimetric immunoassay (HemosIL D-dimer HS 500; Instrumentation Laboratory, Bedford MA, USA; total coefficient of variation < 6.0%) at Rothen Medizinische Laboratorien AG, Basel, Switzerland. Plasma erythritol and xylitol concentrations were quantified by gas chromatography-tandem mass spectrometry (GC-MS/MS) at the Fachhochschule Nordwestschweiz, Muttenz, Switzerland.

Additional analytical and laboratory details are provided in the Supplementary Information.

#### Statistical Analysis

Analyses were performed in Python (v3.10). Full statistical details are provided in the Supplementary Information. Due to insufficient human data on the effects of erythritol on platelet aggregation no *a priori* power calculation was possible. A *posteriori* power calculation indicated that detecting statistically significant differences would have required unrealistically large sample sizes, reflecting more likely the absence of meaningful treatment effects rather than inadequate study design.

The primary endpoint was maximal *ex vivo* platelet aggregation. Maximum aggregation, inclination, lag phase, and de-aggregation were quantified at baseline (t = -60 min) and 60 min after acute oral intake of erythritol (50 g), xylitol (33.5 g), or water (control). Treatment effects were evaluated using change scores (Δ = 60 min − baseline). For each aggregation parameter and agonist, paired within-subject contrasts comparing erythritol *vs.* water and xylitol *vs.* water were tested using paired t-tests. Family-wise error was controlled within each parameter and agonist across the two treatment contrasts using Holm adjustment. As a sensitivity analysis, treatment effects on Δ were additionally evaluated using linear mixed-effects models with subject-specific random intercepts; these models yield estimates equivalent to the paired Δ-contrast approach for two timepoints. Statistical significance was defined as adjusted *P* < 0.05.

For longitudinal markers of platelet activation (P-selectin), endothelial activation (sVCAM-1), and coagulation (D-dimer), time and treatment effects were assessed using linear mixed-effects models with subject-specific random intercepts to account for within-subject correlation across repeated measurements. Fixed effects included time and treatment. Because absence of statistical significance does not imply biological equivalence, equivalence and non-inferiority were evaluated in addition to conventional hypothesis testing using the *Two One-Sided Tests (TOST)* procedure ^17^. Equivalence was concluded when the 90% confidence interval of the paired mean difference lay entirely within pre-specified equivalence margins. Equivalence margins were defined *a priori* based on a reference change value (RCV) framework, providing biologically grounded thresholds for changes exceeding expected within-individual variability. Family-wise error was controlled using Holm or Šidák adjustment, as appropriate.

Plasma concentrations of erythritol and xylitol were analyzed descriptively over time. Molar concentrations were used to calculate metabolic ratios. Time-course data were analyzed using linear mixed-effects models with subject-specific random intercepts and fixed effects for time and treatment. No formal hypothesis testing was performed beyond these models.

### Part B. *In vitro* Experiment: Direct Effects of Erythritol or Xylitol on Platelet Aggregation

#### Study Design and Participants

A separate cohort of 11 healthy, non-smoking participants (18–55 years; BMI <30 kg/m²) was recruited for *in vitro* platelet experiments. Inclusion and exclusion criteria were identical to those described in Part A, except for the BMI criterion. Participants arrived in a fasting state.

#### Laboratory Analyses

For *in vitro* platelet aggregation experiments, blood samples were collected, and PRP and PPP were prepared as described in Part A. The platelet concentration in PRP was adjusted to 250 × 10⁹/L using autologous PPP. The adjusted PRP was then incubated for 30 min at room temperature with erythritol (4.5–270 µM), xylitol (3–300 µM), or vehicle control (0.9 % NaCl) ^11,13^. After pre-incubation, PRPs were maintained in suspension with constant stirring (600 rpm) at 37 °C. Following incubation, platelet aggregation was induced with ADP (2 µM) or TRAP-6 (5 µM) and measured turbidimetrically using an optical aggregometer (APACT 4004, Coultronics).

#### Statistical Analysis

For each agonist (ADP (2 µM) and TRAP-6 (5 µM)), maximum platelet aggregation was summarized as mean ± 95% confidence interval across the tested concentration ranges of erythritol and xylitol. To evaluate whether erythritol or xylitol modifies the concentration-response relationship in a dose-dependent manner, linear mixed-effects models were fitted separately for each agonist, including fixed effects for erythritol and xylitol concentration, and their interaction, with a subject-specific random intercept to account for repeated measurements:

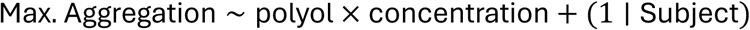

Evidence for differential concentration dependence between erythritol and xylitol was assessed *via* the polyol × concentration interaction term. When appropriate, paired *post-hoc* comparisons between erythritol or xylitol at each concentration level were conducted using Holm-adjusted (or Tukey-adjusted) *P*-values to control the family-wise error rate.

To assess whether erythritol or xylitol induced concentration-dependent deviations from baseline, Dunnett-type contrasts were performed comparing each non-zero concentration with the corresponding 0 µM condition within subjects. These contrasts were evaluated using paired comparisons with Holm adjustment for multiple testing.

## Results

### Part A. Effects of Acute Oral Intake of Erythritol or Xylitol

Initially, 20 healthy participants were enrolled in the study, of which 16 received the allocated intervention. After the exclusion of two participants because of aberrant platelet responses, data from 14 participants (10 women and 4 men; age, 26.5 ± 5.5 years; BMI, 21.5 ± 1.9 kg/m²) were included in the final analysis. Not all participants completed all three study visits; the numbers analyzed for each intervention are provided in Supplemental Figure S1.

### *Ex vivo* Platelet Aggregation

Neither erythritol nor xylitol intake significantly altered *ex vivo* platelet aggregation compared with water across the tested agonists and aggregation parameters (Tables 1 and S2). Effect estimates were generally centered around zero, with small to moderate effect sizes (Cohen’s *dz*, |*dz*| ≤ 0.68), and no between-treatment contrast remained significant after Holm correction for multiple testing. Under stimulation with thrombin receptor-activating peptide-6 (TRAP-6; 5 µM), erythritol was associated with a nominally prolonged lag phase compared with water (unadjusted *P* = 0.049); however, this difference did not remain significant after correction for multiple testing and was not observed with xylitol. No significant treatment effects were detected for any aggregation parameter under adenosine diphosphate (ADP; 2 µM) or higher-concentration TRAP-6 (10 µM) stimulation. Effect estimates were centered around zero across all tested agonists and parameters (Table S2), consistent with the absence of treatment effects.

Results are presented as individual adjusted maximum *ex vivo* platelet aggregation before and 60 min after intake of erythritol, xylitol, or water (Figure 1), and as paired standardized effect sizes (Cohen’s *dz*) in a forest plot (Figure S3). All baseline aggregation values were within the reference ranges, indicating that baseline platelet responsiveness was within the expected physiological range established in healthy volunteers in our routine laboratory according to standardized recommendations ^16,18^. Although a ceiling effect cannot be excluded, particularly with the higher TRAP-6 concentration (10 µM), it is less likely at the lower agonist concentrations (ADP 2 µM and TRAP-6 5 µM), which provide a greater dynamic range for detecting an increase in platelet responsiveness.

**Figure 1.**
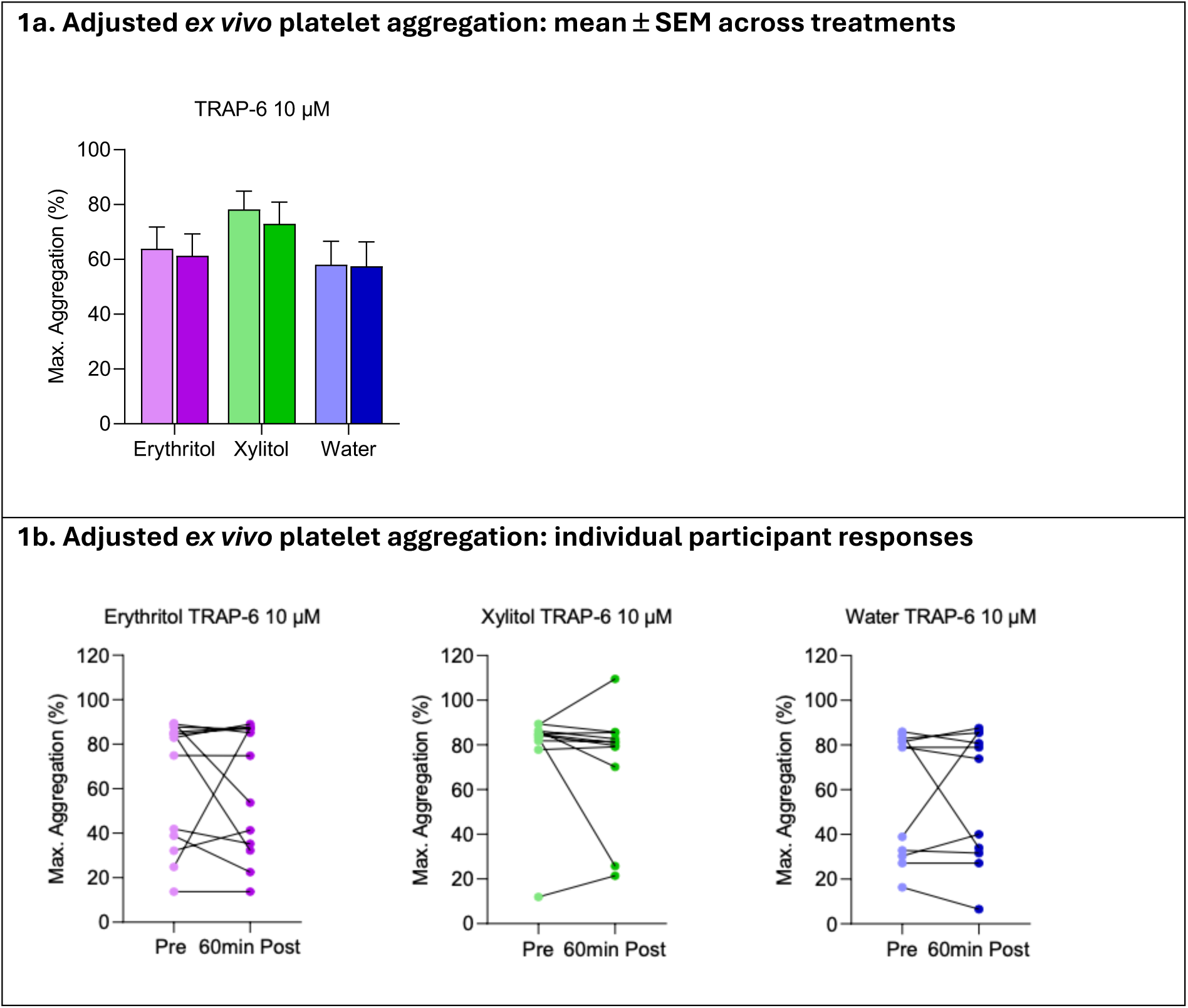
Effects of oral intake of erythritol or xylitol on *ex vivo* platelet aggregation. Adjusted maximum *ex vivo* platelet aggregation (%) in response to TRAP-6 (10 µM) before and 60 min after acute oral intake of erythritol (50 g), xylitol (33.5 g), or water (control). **A)** mean ± SEM across treatments; **B)** individual participant responses. Light-colored bars and circles indicate pre-intake measurements, dark-colored bars and circles indicate post-intake measurements. Statistical comparisons were performed using linear mixed-effect models. Sample sizes: erythritol, n = 13; xylitol and water, n = 11.

### Markers of Platelet Activation, Endothelial Activation, and Coagulation

Neither erythritol nor xylitol significantly affected circulating markers of platelet activation (P-selectin), endothelial activation (sVCAM-1), or coagulation (D-dimer) over time (Figure 2). Observed temporal fluctuations were small and inconsistent, and no between-treatment differences remained significant after correction for multiple testing. Where nominal differences were observed, effect sizes were within the range of biological variability and did not exceed equivalence margins based on Reference Change Values (RCVs). Equivalence testing further showed that post-prandial changes remained within prespecified RCV-based equivalence margins relative to baseline and the water control. Collectively, these findings provide no evidence of acute platelet activation, endothelial activation, or coagulation following erythritol or xylitol intake. Additional results are provided in the Supplementary Information (Tables S4a-f, S5a-f and S6a-f).

**Figure 2.**
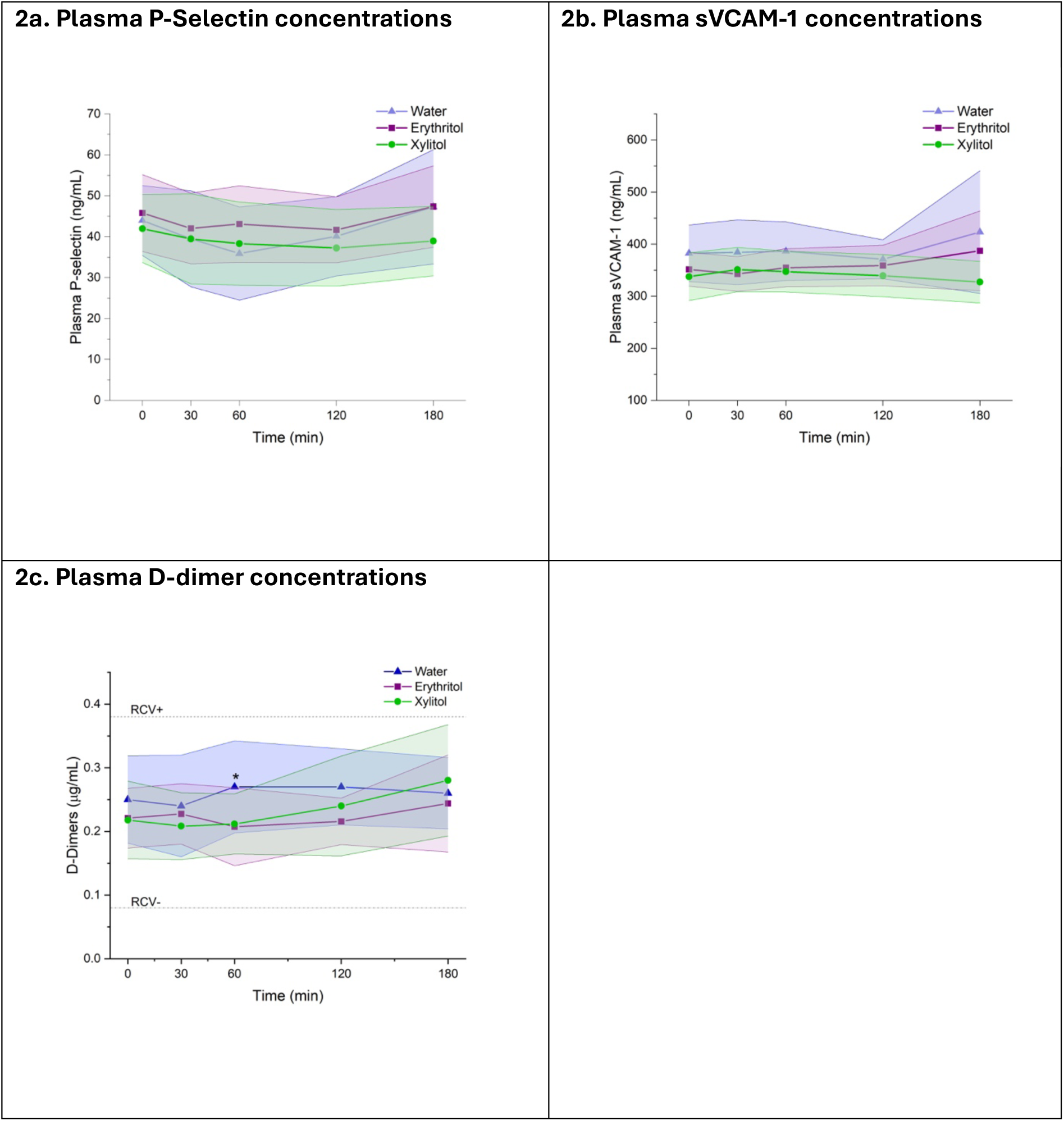
Effects of oral intake of erythritol or xylitol on circulating markers of platelet activation, endothelial activation, and coagulation. Plasma concentrations of **A)** P-selectin, **B)** sVCAM-1, and **C)** D-dimer before and 60 min after acute oral intake of erythritol (50 g), xylitol (33.5 g), or water (control). Data are expressed as mean ± 95% CI. Statistical comparisons were performed using linear mixed-effects models with subject-specific random intercepts and Šidák adjustment. Reference change values (RCV) indicate the expected biological and analytical variability for D-dimer ^19^ relative to which the observed changes are shown. * *P* < 0.05. Sample sizes: A) and B) erythritol, n = 13; xylitol and water, n = 11; C) erythritol, xylitol, and water, n = 11.

### Plasma Erythritol and Xylitol Concentrations

Plasma concentration-time profiles showed a lag of approximately 30 min before peak concentrations of erythritol and xylitol were observed. Oral intake of 50 g erythritol led to pronounced peaks of plasma erythritol, accompanied by smaller but detectable increases in xylitol concentrations (Figure 3a and Table S7). Conversely, oral intake of 33.5 g xylitol produced rapid and pronounced peaks of plasma xylitol, while erythritol concentrations increased more slowly and reached a plateau between 60 and 180 min (Figure 3b), suggesting delayed formation. Estimated area-under-the-curve ratios (AUC₀_–_₁₈₀ _min_ for erythritol-to-xylitol and AUC₀_–_₁₈₀ _min_ for xylitol-to-erythritol) indicated minimal conversion of erythritol to xylitol (0.02%) and substantially greater conversion of xylitol to erythritol (13.7%). After water intake, plasma erythritol and xylitol concentrations remained near the assay’s limit of detection (Figure 3c).

**Figure 3.**
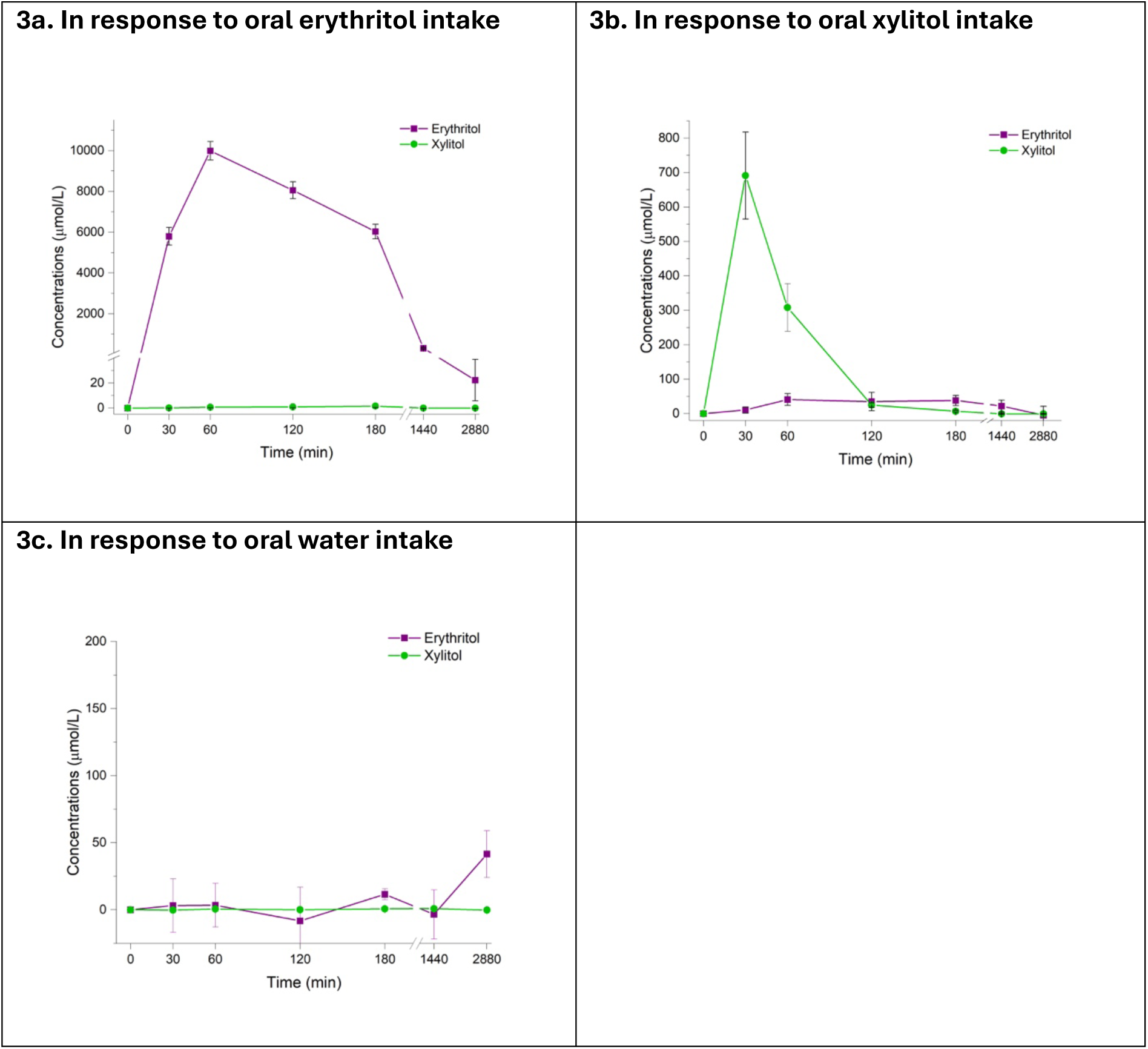
Effects of oral intake of erythritol or xylitol on plasma erythritol and xylitol concentrations. Changes from baseline in plasma erythritol and xylitol concentrations (µmol/L) over 48 h after acute oral intake of **A)** erythritol (50 g), **B)** xylitol (33.5 g), or **C)** water (control). Values represent changes from baseline, calculated by subtracting the fasting concentration from each post-intervention concentration. Data are expressed as mean ± SEM. Sample sizes: erythritol, n = 13; xylitol, and water, n = 11.

### Part B. *In vitro* Experiment: Direct Effects of Erythritol or Xylitol on Platelet Aggregation

Complementary *in vitro* experiments were performed using platelet-rich plasma from an independent cohort of healthy participants to assess concentration-dependent effects of erythritol and xylitol on platelet aggregation under controlled conditions, independent of gastrointestinal absorption, metabolism, and other systemic factors. Data from 11 participants (9 women and 2 men; age, 27.6 ± 3.2 years; BMI 21.7 ± 2.0 kg/m^2^; mean ± SD) were included in the final analysis.

Maximum platelet aggregation was assessed after incubation of PRP with increasing concentrations of erythritol or xylitol, followed by stimulation with ADP (2 µM) or TRAP-6 (5 µM) (Figure 4).

**Figure 4.**
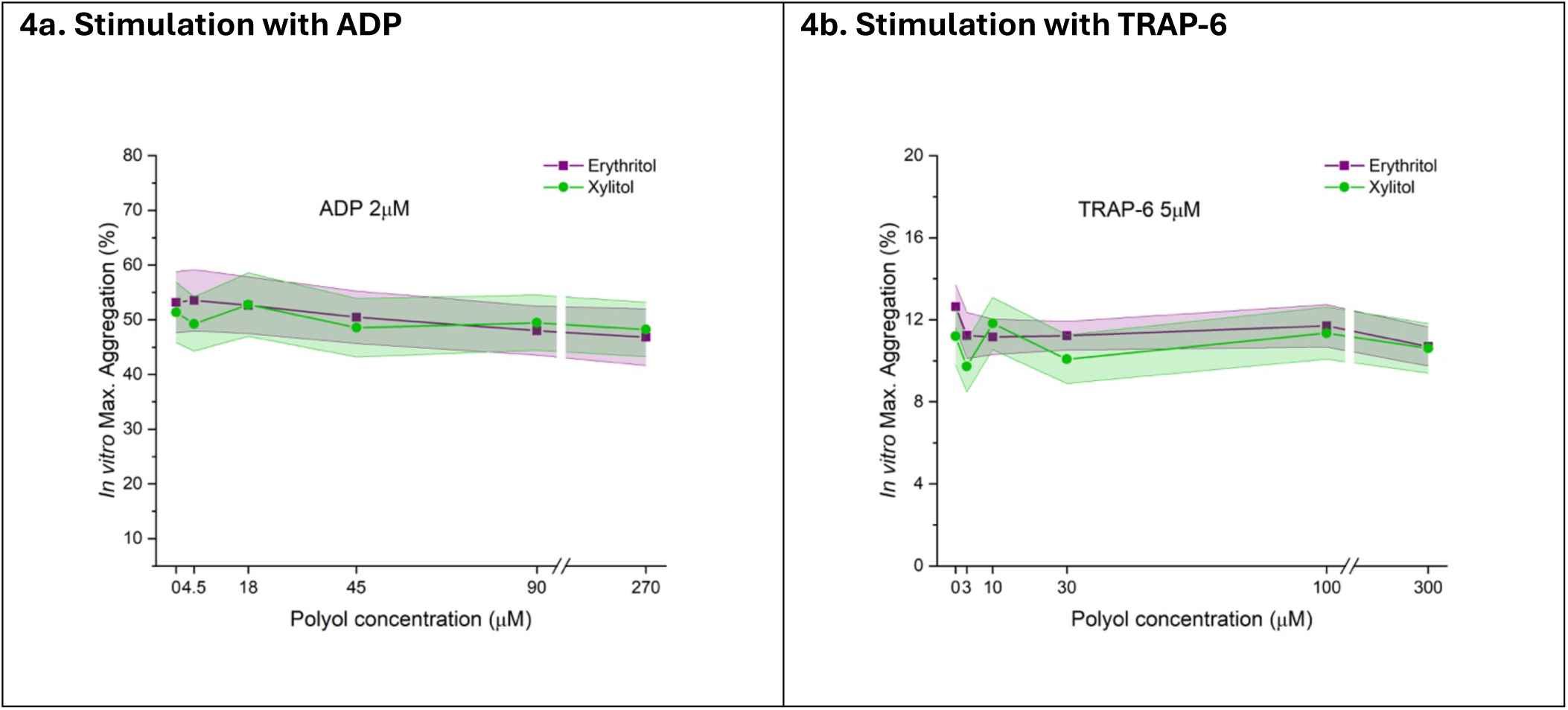
*In vitro* experiment: concentration–response curves of platelet aggregation following erythritol or xylitol incubation. Mean maximum platelet aggregation in platelet-rich plasma following incubation with increasing concentrations of erythritol or xylitol. Panels show aggregation induced with **A)** ADP (2 µM) and **B)** TRAP-6 (5 µM). Points represent mean aggregation values and shaded bands indicate 95% confidence intervals. Sample size: erythritol and xylitol, n = 11.

Wald χ² tests showed no significant overall effects of treatment, concentration, or treatment × concentration interaction on *in vitro* platelet aggregation under either ADP or TRAP-6 stimulation (Table S8a). Within-subject variability at baseline was assessed from paired differences between erythritol₀ and xylitol₀ measurements obtained in the absence of added polyols (Table S8b). Variability was significantly greater under ADP than under TRAP-6 stimulation (F-ratio = 5.81, df = 10,10; *P* = 0.010). Paired differences did not significantly deviate from normality for either ADP (Shapiro-Wilk W = 0.96, *P* = 0.758) or TRAP-6 (W = 0.90, *P* = 0.205), supporting the variance comparison (Table S8b). The greater within subject variability under ADP stimulation resulted in wider confidence intervals and reduced precision for equivalence testing, despite the absence of systematic changes in mean aggregation across erythritol and xylitol concentrations (Table S8b).

Equivalence to vehicle control was assessed using the *Two One-Sided Tests* (TOST) procedure ^17,20^, with pre-specified equivalence bounds of ± 5 percentage points (Table S8c). For ADP-induced aggregation, equivalence could not be established at any concentration because the 90% confidence intervals extended beyond the pre-specified equivalence bounds and greater within-subject variability. In contrast, TRAP-6-induced aggregation following exposure to either erythritol or xylitol was equivalent to vehicle control across all tested concentrations, with the corresponding 90% confidence intervals contained within the equivalence bounds (Table S8c). Family-wise error was controlled using Holm adjustment ^21^.

Figure 5 displays the full distribution of individual responses and illustrates the greater interindividual variability observed with ADP stimulation. Despite this variability, there was no systematic concentration-dependent shift in platelet aggregation with either erythritol or xylitol. Consistent with the formal statistical analyses, no concentration-dependent effect of neither erythritol nor xylitol was detected under ADP or TRAP-6 stimulation (Table S8d).

**Figure 5.**
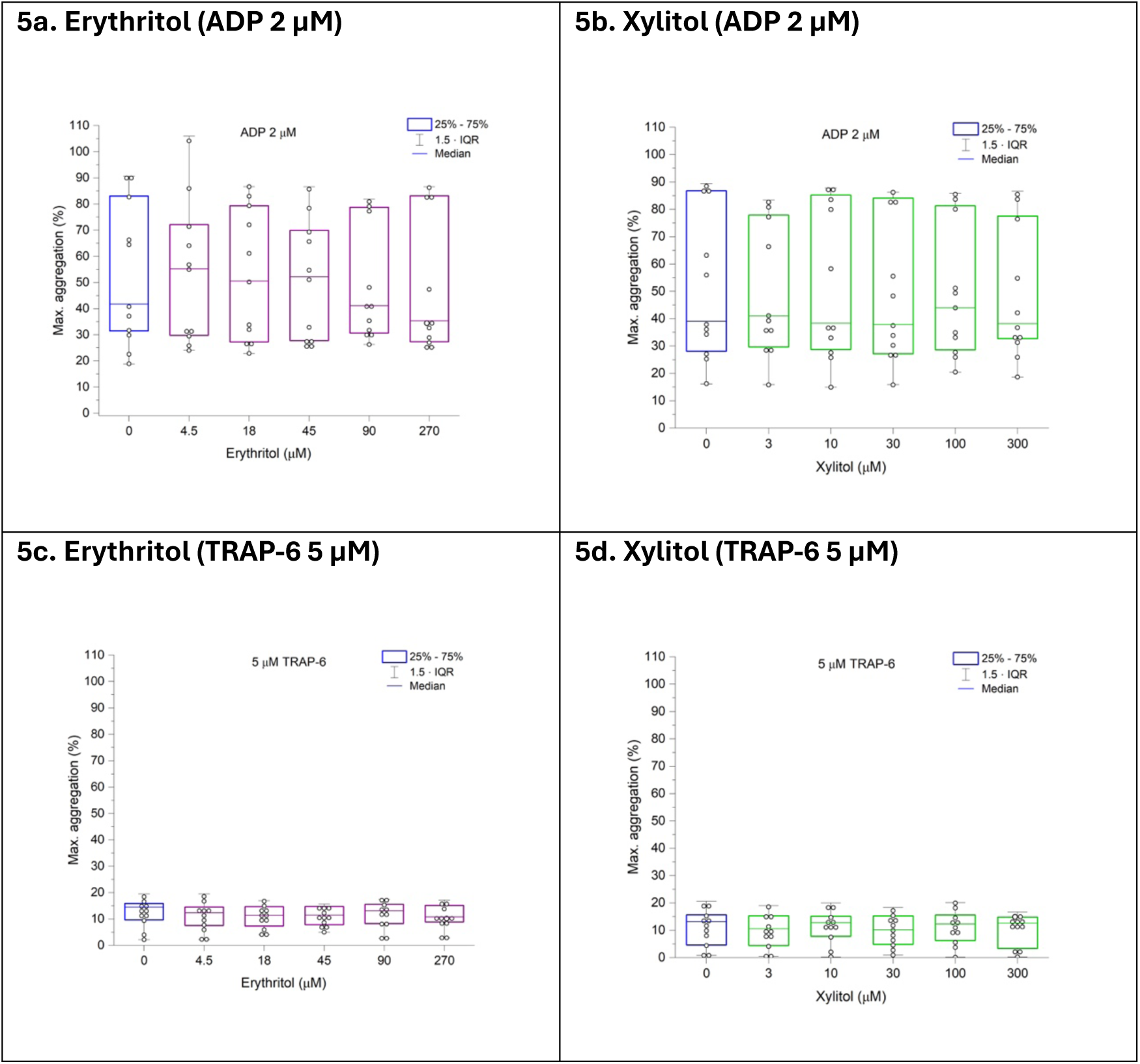
*In vitro* experiment: individual participant platelet aggregation responses following erythritol or xylitol incubation. Individual maximum platelet aggregation in platelet-rich plasma following incubation with increasing concentrations of erythritol or xylitol. Panels show **A)** erythritol + ADP (2 µM), **B)** xylitol + ADP (2 µM), **C)** erythritol + TRAP-6 (5 µM), and **D)** xylitol + TRAP-6 (5 µM). Aggregation was calculated from changes in light transmission using autologous platelet-rich plasma (PRP) and platelet-poor plasma (PPP) as the 0% and 100% reference points, respectively. Accordingly, values slightly exceeding 100% may occur when light transmission in an aggregated PRP sample exceeds the PPP reference value and do not indicate aggregation of more than 100% of platelets. The variability observed following stimulation with 2 µM ADP reflects interindividual differences in platelet responsiveness under submaximal agonist stimulation. Box plots indicate the median (center line), interquartile range (25^th^-75^th^ percentiles), and whiskers extending to 1.5× the interquartile range; points represent individual donors; blue boxes indicate incubation with vehicle control, while purple and green boxes indicate incubation with erythritol or xylitol, respectively. Samples size: erythritol and xylitol, n = 11.

## Discussion

In this randomized crossover study of healthy participants, acute oral intake of erythritol or xylitol did not increase *ex vivo* platelet aggregation compared with water. Consistent with these findings, neither erythritol nor xylitol induced relevant changes in circulating markers of platelet activation, endothelial activation, or coagulation. Complementary *in vitro* experiments further showed no concentration-dependent increase in platelet aggregation following direct exposure to erythritol or xylitol. Taken together, these findings provide no evidence that acute erythritol or xylitol exposure enhances platelet reactivity or promotes a prothrombotic state under the conditions studied.

These findings are relevant in the context to reduce the cardiometabolic burden associated with excessive intake of added sugars. Although reducing overall sugar intake remains the primary preventive strategy, partial replacement of sugar with alternative sweeteners with a more favorable metabolic profile may represent a complementary approach. Erythritol and xylitol have attracted particular interest because they provide sweetness with a significantly lower caloric load than sucrose, minimal glycemic and insulinemic responses, and the ability to stimulate the release of gastrointestinal satiation hormones such as glucagon-like peptide-1 (GLP-1), peptide YY (PYY), and cholecystokinin (CCK), making them particular interesting in the context of metabolic health ^9^.

Observational studies have raised safety concerns by reporting associations between elevated plasma erythritol and xylitol concentrations and increased risk of cardiovascular disease (CVD), and major adverse cardiovascular events (MACE) ^11,13^. However, these studies did not assess actual dietary intake of erythritol or xylitol, limiting causal interpretation. Elevated plasma erythritol or xylitol concentrations may instead result from endogenous production, which is known to increase in various chronic metabolic conditions and may primarily reflect underlying metabolic dysregulation rather than a causal role of sweetener intake ^22^. Supporting this notion, in 2019 Wang *et al.* observed higher erythritol concentrations associated with CVD risk in samples collected before erythritol’s approval as a food additive by the Food and Drug Administration (FDA) in 2001, indicating endogenous synthesis ^23^. Consistent with this interpretation, Mendelian randomization analyses found no evidence of a causal link between erythritol and cardiometabolic traits or T2DM ^24^. Mechanistically, chronic hyperglycemia may contribute to increased endogenous polyol production *via* upregulation of the pentose phosphate and polyol pathways, offering a plausible explanation for the association between elevated erythritol concentrations, T2DM and cardiovascular risk ^7^.

Direct vascular effects of erythritol and xylitol in humans have been investigated in a limited number of clinical studies. Flint *et al.* reported that in participants with T2DM, acute intake of 24 g erythritol improved small vessel endothelial function two hours post-intake, while daily intake of 36 g over four weeks reduced central aortic stiffness ^25^. In contrast, Bordier *et al*. found that daily supplementation with 36 g erythritol or 24 g xylitol for five weeks did not significantly affect vascular function, as assessed by left brachial pulse wave velocity and retinal vessel diameter, in normoglycemic adults with obesity ^26^. Taken together, these findings suggest that the intake of erythritol or xylitol may exert either neutral or potentially protective effects on endothelial function, with outcomes possibly influenced by study population and underlying metabolic and glycemic status. Nevertheless, in two recent interventional studies, a direct effect of dietary erythritol and xylitol intake on *ex vivo* platelet aggregation in healthy humans has been described: in a parallel study with ten healthy participants, acute oral intake of 30 g of erythritol led to a significant increase in maximum platelet aggregation (using ADP or TRAP-6 as agonist) 30 min after intake, but not after 30 g glucose intake ^14^. In a separate study by the same group, acute oral intake of 30 g xylitol also resulted in a significant increase in maximum platelet aggregation 30 min post-intake ^13^. However, the study design in those two separate studies included three separate study populations, each receiving only one of the test solutions, thereby preventing direct within-subject comparisons ^13,14^. Moreover, the studies lacked a control arm, which is particularly important when studying platelet aggregation, given the individual variability and natural fluctuations in platelet aggregation, which is sensitive to various stimuli and can be influenced by food intake or physical activity ^27–29^.

Our findings differ from those of Witkowski *et al.* ^13,14^. In this randomized crossover study, neither erythritol nor xylitol affected *ex vivo* platelet aggregation or induced relevant changes in circulating markers of platelet activation (P-selectin), endothelial activation (sVCAM-1), or coagulation (D-dimer) in healthy participants. Whereas Witkowski *et al*. reported increased platelet aggregation across participants, we observed interindividual variability within the expected biological range ^13,14^. Several methodological differences should be considered when comparing the studies. First, we assessed platelet aggregation at baseline and 60 min after oral intake, whereas Witkowski *et al*. assessed aggregation at baseline and 30 min. The 60-minute timepoint was selected *a priori* based on previous findings from our group showing peak plasma erythritol-concentrations 60 min after oral intake.^30^ Consistent with these findings, erythritol concentrations in the present study peaked at 60 min, whereas xylitol concentrations peaked at 30 min. Because platelet aggregation was not assessed at 30 min, we cannot exclude an effect restricted to an earlier time window, particularly for xylitol. However, P-selectin, sVCAM-1, and D-dimer measured at 30 min showed no increase, providing no evidence of concomitant platelet, endothelial, or coagulation activation at this time point. Second, the administered erythritol dose was higher in our study than in that of Witkowski *et al.* (50 g vs. 30 g), whereas the xylitol doses were similar (33.5 g vs. 30 g). The higher erythritol dose would be expected to increase the ability to detect an acute effect attributable to erythritol. Finally, unlike Witkowski *et al.*, we did not perform a complete dose-response analysis. Instead, agonist concentrations were selected *a priori* within the ranges in which Witkowski *et al.* reported enhanced platelet responsiveness (ADP 2 µM; TRAP-6 5 and 10 µM). The lower agonist concentrations (ADP 2 µM and TRAP-6 5 µM) provided sufficient dynamic range to detect an increase in platelet responsiveness, while the higher TRAP-6 concentration (10 µM) allowed assessment under stronger stimulation. Thus, although differences in experimental design should be considered when comparing the studies, our protocol included conditions under which an erythritol- or xylitol-associated increase in platelet responsiveness could have been detected.

Plasma concentrations of P-selectin, a key marker of platelet degranulation and thrombo-inflammatory activity ^31^, and sVCAM-1, a biomarker of endothelial activation and dysfunction ^32,33^, are also influenced by multiple factors, including physical activity, genetics, sex, smoking, ethnicity, blood pressure, and circadian variation ^34–40^. In our study, all concentrations observed were within the normal range ^41–48^, and did not differ significantly between treatments or across timepoints. Minor fluctuations over the 180 min period likely reflect physiological variability. D-dimer, a marker of fibrinolysis and thrombus formation, likewise stayed far below clinical decision limits (typically < 500 ng/mL [0.5 µg/mL]), confirming the absence of any pro-thrombotic effect ^49^. A transient decrease at 60 min after erythritol and xylitol intake reached nominal statistical significance compared with water, but the absolute differences were minimal (ca. 0.02-0.04 µg/mL) and far smaller than the Reference Change Value (RCV; 0.08-0.38 µg/mL). Equivalence testing (TOST) confirmed that 90% confidence intervals for treatment differences were contained within the predefined equivalence bounds (± 0.10 µg/mL), supporting the absence of clinically meaningful changes.

By assessing plasma concentrations of erythritol or xylitol in the present study, we confirmed previous findings that erythritol is rapidly absorbed, reaching peak plasma concentrations approximately 60 min after intake, and remains mildly elevated for up to 48 hours post-intake ^11,30^. Although methodological differences in erythritol quantification may result in variations in absolute values, overall concentration profiles appear comparable across studies, with a consistent kinetic pattern ^11^. Only a few studies have assessed plasma xylitol concentrations after intake, and absolute values may vary depending on the analytical method used. Nevertheless, our findings align with previous research indicating that peak xylitol concentrations occur 30 min after oral intake and return to baseline concentrations within 4-6 hours, or by 24 hours at the latest ^13^.

In addition to the *ex vivo* platelet aggregation assessments following acute oral intake of erythritol, xylitol or water, we performed complementary *in vitro* experiments in which platelets were directly exposed to erythritol or xylitol across a range of concentrations to assess potential direct effects. Because elevated glucose concentrations can enhance platelet aggregation *in vitro,* possibly due to increased osmolarity ^50^, a similar effect of erythritol and xylitol was considered plausible. However, direct exposure to erythritol or xylitol did not alter platelet aggregation under either ADP or TRAP-6 stimulation, failing to reproduce earlier findings ^11,13^.

Although interindividual variability was comparable between agonists, intra-subject variability was lower for TRAP-6 than for ADP-induced aggregation, likely reflecting differences in activation mechanisms. Nevertheless, no systematic concentration-dependent effects of erythritol or xylitol were observed *in vitro*, consistent with the absence of effects observed *ex vivo*.

This study has several strengths. First, the randomized crossover design allowed each participant to serve as their own control, and the inclusion of an additional control arm further strengthened the comparisons. Second, all procedures related to study visits, blood collection, and subsequent measurements were standardized and conducted according to established recommendations for assessing of platelet function by light transmission aggregometry ^16^, ensuring the robustness of the findings. Third, the administered doses of erythritol (50 g) and xylitol (33.5 g) were relatively high, increasing the likelihood of detecting measurable effects. Fourth, by combining *ex vivo* and *in vitro* platelet aggregation assessments, the study examined platelet responses from both physiological and mechanistic perspectives. Fifth, quantification of erythritol and xylitol concentrations enabled correlation analyses. Finally, the 48-hour measurement period provided an extensive overview of erythritol and xylitol kinetics following intake.

Nonetheless, this study also has some limitations. First, although the sample size was modest, the randomized crossover design minimized between-subject variability and enabled within-subject comparisons across treatments. Second, participants were predominantly female, young, healthy, and lean, and recruitment did not use prespecified targets to ensure broader demographic representation. This limits the generalizability of our findings, particularly to older individuals and populations with cardiometabolic disease, and precludes meaningful sex-specific analyses. Third, participants could easily identify the water control, whereas the erythritol- and xylitol-containing beverages were indistinguishable. Thus, although investigators and laboratory personnel were fully blinded to treatment allocation, participant blinding was incomplete. Fourth, maximum platelet aggregation was assessed at a single post-intake time point (60 min), limiting our ability to detect potentially transient effects at earlier or later time points. This time point coincided with peak plasma erythritol concentrations, whereas xylitol concentrations peaked at 30 min. Finally, the in vitro experiments included a negative control (sodium chloride, NaCl) but no positive control. Although platelet responsiveness to ADP and TRAP-6 was readily detectable, the absence of a positive control specifically demonstrating enhanced platelet aggregation limits the interpretation of the negative findings.

In conclusion, reducing the overall intake of added sugars remains a key strategy for promoting cardiovascular health. In healthy participants, acute oral intake of the alternative sweeteners erythritol (50 g) or xylitol (33.5 g) did not increase *ex vivo* platelet aggregation or circulating markers of platelet activation (P-selectin), endothelial activation (sVCAM-1), or coagulation (D-dimer) compared with water. These findings do not support an acute prothrombotic effect of erythritol or xylitol under the conditions studied. However, given the modest sample size, short exposure period, and inclusion of healthy participants, these results should not be extrapolated to long-term cardiovascular safety. Larger, longer-term studies, particularly in individuals at increased cardiovascular risk, are needed to determine the clinical implications of sustained erythritol and xylitol consumption.

## Data Availability

The data that support the findings of this study are available from the corresponding author upon reasonable request.

## Acknowledgements

We gratefully acknowledge all participants for their contribution to this study. We thank Ann-Kathrin Draxler, Veronika Walhöfer and Geena Spielmann for their assistance. The authors used ChatGPT (OpenAI, 2025) and Grammarly to assist with language editing and improving readability. The authors reviewed and edited the output and take full responsibility for the content of this publication.

## Author Contributions

Conceptualization, B.K.W. and A.C.M.-G.; Methodology, B.K.W., A.C.M.-G., and A.A.-S.; Formal analysis, B.K.W., A.A., E.F., M.T., T.K., J.D., S.G., A.A.-S., and A.C.M.-G.; Investigation, A.A., E.F., T.K., and M.T.; Resources, B.K.W., A.C.M.-G., and A.A.-S.; Data curation, B.K.W., A.A., E.F., M.T., T.K., J.D., A.A.-S., and A.C.M.-G.; Writing - original draft, B.K.W., A.A., and A.C.M.-G.; Writing - review and editing, all authors; Visualization, B.K.W., A.A., J.D., and A.C.M.-G.; Supervision, B.K.W., A.A.-S., and A.C.M.-G.; Project administration, B.K.W., A.A.-S., and A.C.M.-G.; Funding acquisition, B.K.W., A.C.M.-G, and A.A.-S.

## Supplemental Material

Supplementary Methods

Supplementary Results

Supplementary Figures and Tables S1- S8d

## Non-standard Abbreviations and Acronyms

ADP: adenosine diphosphate
CVD: cardiovascular disease
T2DM: type 2 diabetes mellitus
TRAP-6: thrombin receptor-activating peptide-6
TOST: Two One-Sided Tests
RCV: Reference Change Value
PPP: platelet-poor plasma
PRP: platelet-rich plasma
GLP-1: glucagon-like peptide-1
PYY: peptide YY (PYY)
CCK: cholecystokinin
MACE: major adverse cardiovascular events
ELISA: enzyme-linked immunosorbent assay
GC-MS/MS: gas chromatography-tandem mass spectrometry
PFA: platelet function assay

---

**Table 1:**
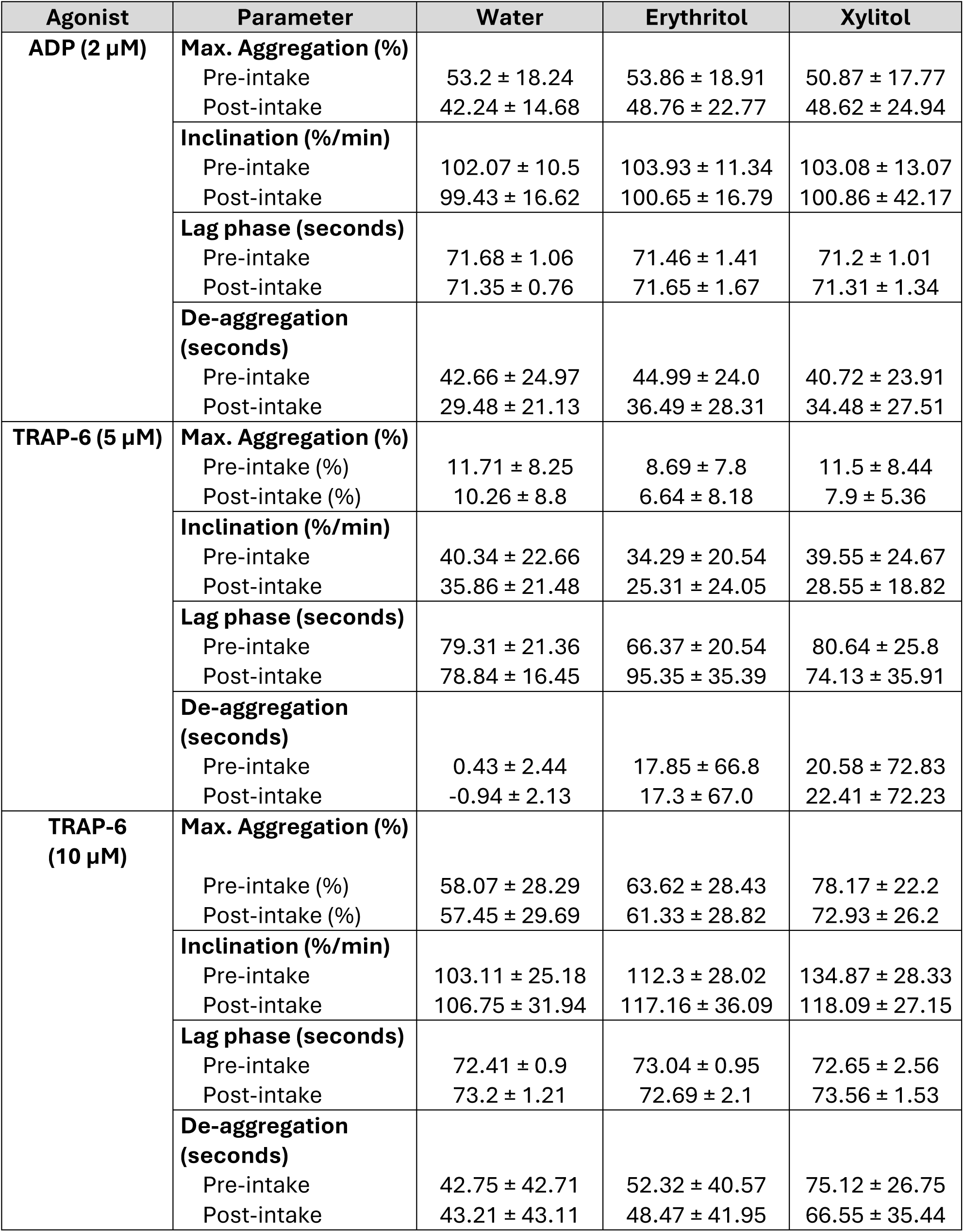
Mean (± SD) *ex vivo* Platelet Aggregation Parameters under ADP (2 µM) and TRAP-6 (5 and 10 µM) Stimulation in Response to Acute Oral Intake of Erythritol, Xylitol, or Water.

